# Benefits of local consolidative treatment in oligometastases of solid cancers: a stepwise-hierarchical pooled analysis and systematic review

**DOI:** 10.1101/2020.08.05.20168724

**Authors:** Chai Hong Rim, In-Soo Shin, Sunmin Park, Hye Yoon Lee

## Abstract

**Purpose:** Any available evidence regarding the application of local consolidative therapy (LCT) for oligometastases is from phase 2 and observational studies. This study aimed to evaluate the oncologic benefits of LCT in oligometastatic setting.

**Methods:** The MEDLINE, EMBASE, and Cochrane library were searched. We applied stepwise analyses that enabled the evaluation of data from randomized controlled trials (RCTs), balanced studies (e.g. without significant differences regarding major prognosticators between arms), and all studies separately and in a hierarchical manner

**Results:** Thirty-one studies including seven randomized trials were reviewed. Pooled analyses of the effect of LCT on overall survival (OS) revealed odds ratios (ORs) of 3.04 (95% confidence interval [CI]: 2.28–4.06, p<0.001), 2.56 (95% CI: 1.79–3.66, p<0.001), and 1.41 (95% CI: 1.02–1.95, p=0.041) for all studies, balanced studies, and RCTs, respectively. The corresponding ORs for progression-free survival were 2.82 (95% CI: 1.96– 4.06, p<0.001), 2.32 (95% CI: 1.60–3.38, p<0·001), and 1.39 (95% CI: 1.09–1.80, p=0.009), respectively. The benefit of LCT was higher in non-small cell lung cancer (OR: 3.14, p<0.001; pooled 2-year OS: 65.2% vs. 37.0%) and colorectal cancer (OR: 4.11, p=0.066; pooled two-year OS: 66.2% vs. 33.2%) than in prostate (OR: 1.87, p=0.006; pooled three-year OS: 95.6% vs. 92.6%) and small cell lung cancer (OR: 1.04, p=0.942; pooled one-year OS: 60.7% vs. 42.8%). Complications were generally mild.

**Conclusion:** LCT provides oncologic benefits in the oligometastatic setting, although such benefits were less evident in RCTs than in data from observational studies. The appropriate LCTs should be carefully selected, considering their feasibility and disease types.

## Introduction

To date, cancer treatments have been prescribed based on the pathologic stage of progression. The highest solid cancer stage indicates a systemic disease that has spread beyond the primary tumor and lymphatics, and has little-to-no chance of being cured. Systemic administration of chemotherapy is regarded as the only valid option, while local modalities such as surgery or radiotherapy are deemed ineffective in terms of survival.

However, long-term survival is not uncommon among patients with metastases who have undergone successful local salvage. In the late 20^th^ century, a pivotal case series revealed that patients who underwent resection of hepatic metastases from colorectal cancer had five-year survival rates of 28%–37% (1-3); this rate reached 58% in a more recent series.(4) An International Registry of Lung Metastases study revealed five- and ten-year survival rates of 36% and 26%, respectively, after curative resection of lung metastases.(5) Survival outcomes were affected by lower metastatic burdens or tumor markers, which pointed to the gradually progressing nature of the metastatic cascade and presence of an intermediate state, i.e., *oligometastasis*.

Nevertheless, more than two-thirds of such patients ultimately experience polymetastases, and open surgery might be burdensome for some patients whose chance of cure is uncertain and who are debilitated by their cancer. Meanwhile, the practical and clinical consideration of oligometastases has increased with technological advances in radiotherapy. Given the development of conformal technologies based on computed tomography planning, such as stereotactic body radiotherapy (SBRT), non-invasive and ablative irradiation methods for metastatic lesions have become feasible.(6)

Extensive literature has recently emerged regarding the application of local consolidative treatment (LCT) for oligometastases (7, 8); however, the vast majority publications describe single-arm observational studies. This is partly because it can be difficult to design randomized controlled trials (RCTs) involving patients with metastases given the ethical considerations and patients’ widely varying clinical characteristics. The biological understanding of oligometastatic disease has evolved but remains unclear. Therefore, whether patients can benefit from local treatment to their metastases, and whether “oligometastases” even exists as a status, remain controversial. (9, 10)

The aim of this meta-analysis was to assess the efficacy of LCT for patients with oligometastases arising from any type of solid cancers, thereby validating the benefit of LCT and helping clinical decision.

## Methods

### Study protocol

Our study adhered to the Preferred Reporting Items for Systematic Reviews and Meta-Analysis (PRISMA) guidelines. The meta-analysis was designed to answer the following PICO question: “*Does LCT confer an oncologic benefit for patients with oligometastases?*” By implication, the response to this question would demonstrate whether a clinically meaningful “oligometastatic” status exists. LCT was defined as any local treatment targeted toward metastases and/or remnant primary disease in an oligometastatic setting. The MEDLINE, EMBASE, and Cochrane library were systematically searched by two independent reviewers for articles published up to March 4, 2020. The following search terms were used: (oligometastasis OR oligometastases OR oligometastatic OR “limited metastatic” OR “limited metastasis” OR “limited metastases”) AND survival AND (randomised OR randomized OR versus OR comparison OR compare OR controlled) with no language restrictions. The reference lists of the extracted articles were also searched. The retrieved published studies compared the LCT and control arms. Studies published before the year 2000 were excluded to avoid introducing potential bias from outdated treatments. Online registration of the protocol was not performed.

### Selection criteria

The inclusion criteria were as follows: 1) controlled trial involving patients with oligometastases that compared the outcomes of those who underwent LCT versus a control group; 2) 10 or more patients in each arm; 3) at least one primary endpoint provided; and 4) oligometastases defined as five or fewer metastases or as metastases that could definitely be encompassed and treated with LCT. The primary endpoints were overall survival (OS) and progression-free survival (PFS). Grade ≥3 complications related to LCTs were assessed subjectively. For multiple studies from a single institution, only those with the larger number of patients and no (or negligible) overlapping patient pools were all included. Duplicate studies and those with irrelevant formats (e.g., reviews, editorials, letters, or case reports) were automatically filtered. Full-text reviews were performed to identify studies that fulfilled the inclusion criteria.

### Data extraction and quality assessment

Data were extracted using a pre-standardized form; PFS and OS data were estimated from descriptive graphs in the absence of numerical reports. Quality assessment was performed using the Newcastle-Ottawa Scale (11) for cohort studies. Among the three scale domains (“selection” [four points], “comparability” [two points], and “outcome” [three points]), the difference in scores among the studies was mostly due to “comparability.” To avoid subjectivity, we defined the rationale for evaluating comparability based on discussions between clinical oncologists and a biostatistician as follows: 1) RCTs were assigned a full score (two points) unless they had serious clinical differences between comparison arms or flaws in their study designs; 2) statistically matched cohorts (e.g., propensity score matching) or cohorts without significant differences in major clinical indicators were assigned one point; and 3) those with no statistical comparisons or no possibility of clinically significant differences between arms were allotted zero points. Major clinical indicators included the number of metastases, performance status, age, T stage, N stage, prostate-specific antigen (for prostate cancer), and primary disease control; the locations of the metastases were not considered. Studies that scored eight points or higher were considered high quality and balanced, those with six or seven points were medium quality, and lower scores were indicative of low quality.

### Statistical analysis

Pooled analyses of primary endpoints were performed (considering the study quality) in a stepwise hierarchical manner. Overall analysis of all the studies were first performed; next, pooled analyses of balanced studies (≥8 points on the Newcastle-Ottawa scale) were performed, followed by pooled analyses of only the RCTs. Considering the varying study designs, treatment modalities, and clinical characteristics, the random effects model was used for the first two analyses while the fixed effects model was used for the pooled analyses of RCTs. The two-year OS and PFS underwent pooled analysis: the one-year rate was considered when the survival interval was too short or the two-year rate neared 0% (e.g., patients with small cell lung cancer [SCLC] and hepatocellular carcinoma [HCC]); the three- or five-year rates were considered if the survival rates were too high at one or two years (e.g., patients with prostate cancer). Pooled analyses were also performed for studies categorized by specific malignancies using a random effects model. Heterogeneities were assessed using Cochran Q (12) and I^2^ statistics.(13) Significant heterogeneity was considered present when p<0.1 and I^2^ 50%; I^2^ values of 25%, 50%, and 75% corresponded to low, moderate, and high degrees of heterogeneity, respectively. Publication bias was evaluated using funnel plots as well as quantitatively using Egger’s test.(14) If a significant possibility of bias was detected (two-tailed p<0.1),(14) Duval and Tweedie’s trim and fill method (15) was used for sensitivity analysis. Pooled temporal analyses of numerical OS and PFS rates according to cancer type were performed using the Q-test based on analysis of variance. Publication bias assessment was performed only for pooled analyses that included 10 or more studies. All the statistical analyses were performed using the Comprehensive Meta-Analysis software version 3 (Biostat Inc., Englewood, NJ, USA).

## Results

The study included 31 controlled studies (23 retrospective and eight prospective) (9, 16-41) extracted from 1,436 initially searched records across three databases, including 4,762 patients of whom 2,186 and 2,576 were divided into the LCT and control arms, respectively. The study inclusion process is depicted in Figure 1. Eight studies reported conflicts of interests with industrial sponsorship; the remainder had nothing to disclose. Seven studies were RCTs, eight used propensity score matching, 12 reported statistical comparisons between arms involving major clinical indicators, and five had no comparative statistical data. Twelve studies included patients with non-small cell lung cancer (NSCLC), two included patients with SCLC, six included patients with prostate cancer, three included patients with colorectal cancer, two included patients with esophageal cancer, two included patients with HCC, and one each included patients with bile duct, head and neck, sarcoma, and multiple cancers. Most studies (25, 81%) included patients with synchronous and/or metachronous oligometastases, and six (19%) targeted those with metachronous oligometastases. Eleven studies (35%) defined oligometastases as having ≤5 metastases, eight (26%) defined it as having ≤3 metastases, and the remainder used varying definitions (Table 1).

**Table 1.** General information from the included studies

| Author,<br>target<br>disease | Affiliation | Publication | Patient<br>recruit | Study<br>type | LCT group<br>compared to<br>control | Total<br>No. of<br>patients | NOS<br>score | Type of<br>oligometastases;<br>Preceding Tx.<br>For primary dz. | Defined No.<br>of oligomets. | Conflicts<br>of interest |
| --- | --- | --- | --- | --- | --- | --- | --- | --- | --- | --- |
|  |  | Year |  |  |  |  |  |  |  |  |
| He,<br>NSCLC | Sun Yat-sen<br>University,<br>China. | 2017 | 2003–<br>2013 | R | N/A | 21 | 7 | Synchronous and<br>metachronous;<br>OP | ≤3, in lung | None |
| Iyengar,<br>NSCLC | University of<br>Texas<br>Southwestern,<br>US | 2017 | 2014–<br>2016 | P | RCT | 29 | 9 | Synchronous;<br>PR or SD after<br>CTx. | Up to 6 lesions<br>(including<br>primary) in 3<br>organs | None |
| Sheu,<br>NSCLC | MDACC, US | 2014 | 1998–<br>2012 | R | PSM,<br>balanced<br>except higher<br>age | 74 | 9 | Synchronous;<br>no PD after CTx. | ≤3 | None |
| Yano,<br>NSCLC | Kyushu<br>University,<br>Japan | 2010 | 1994–<br>2004 | R | N/A | 93 | 7 | Metachronous;<br>surgery | Controllable<br>with surgery<br>or RTx | None |
| Frost,<br>NSCLC | Charité, Evangelische<br>Lungenklinik, DRK<br>Klinikum Berlin-Mitte, Germany. | 2018 | 2000-2016 | R | PSM | 180 | 9 | Synchronous | 1–4 in one<br>organ | None |
| Gomez,<br>NSCLC | MDACC, London<br>health center,<br>University of<br>Colorado, US & UK | 2019 | 2012–<br>2016 | P | RCT | 49 | 9 | Synchronous and<br>metachronous;<br>CTx. | ≤3 | None |
| Gray,<br>NSCLC | Harvard Medical<br>School, US | 2014 | 2000–<br>2011 | R | younger age<br>(p=0.027) | 66 | 7 | Synchronous | ≤4, brain<br>only | Industrial |
| Hu,<br>NSCLC | Shanghai<br>Jiaotong<br>University,<br>China. | 2019 | 2010–<br>2016 | R | more brain<br>mets, less lung<br>mets.<br>(P<0.001) | 231 | 8 | Synchronous;<br>TKI | ≤5 in single<br>organ | None |
| Song,<br>NSCLC | Cancer Hospital of<br>China Medical<br>University,<br>Liaoning Cancer<br>Hospital and<br>Institute<br>. | 2020 | 2005–<br>2019 | R | PSM, more<br>peripheral<br>location of<br>mets.<br>(p=0.048) | 70 | 9 | Synchronous | ≤5 | None |
| Xu Q,<br>NSCLC | Tongji<br>University,<br>China | 2018 | 2010–<br>2016 | R | Lower T and<br>N stage | 90 | 7 | Synchronous;<br>PR or SD after<br>TKI | ≤5 | None |
| Ni,<br>NSCLC | Shandong First<br>Medical<br>University,<br>China. | 2020 | 2015–<br>2018 | R | no significant<br>difference | 86 | 8 | Synchronous | ≤5 | None |
| Shang,<br>NSCLC<br>(postop) | Shandong<br>University,<br>China. | 2019 | 2005–<br>2016 | R | no significant<br>difference<br>except mets.<br>location | 152 | 8 | Synchronous | ≤5 | None |
| Gore,<br>SCLC<br>(extended) | 57 centers | 2017 | 2010–<br>2015 | P | RCT, more<br>old age in<br>control,<br>p=0.03) | 86 | 9 | Synchronous;<br>PR or CR after<br>CTx. | ≤4 | Industrial |
| Xu<br>SCLC<br>(extended) | Tianjin Medical<br>University,<br>China | 2017 | 2010–<br>2015 | R | PSM, more<br>weight loss<br>patient | 44 | 9 | Synchronous | in one organ or<br>in single RT<br>portal | None |
| Bouman-Wammes,<br>prostate | VUMC,<br>Netherland | 2017 | 2009–<br>2015 | R | higher PSA at<br>Dx. (p=0.015),<br>more single<br>mets (p=0.003) | 63 | 7 | Metachronous;<br>prostatectomy or<br>RTx. | ≤3 | Industrial |
| Lan,<br>prostate | Lanzhou General<br>Hospital of<br>Lanzhou<br>Command,<br>China. | 2019 | 2005–<br>2016 | R | lower PSA<br>(p=0.003), cT (p<br><0.001), N stage<br>(p=0.015), fewer<br>bone mets<br>(p=0.019) | 111 | 7 | Synchronous | ≤5 | None |
| Ost, prostate | Six institutions in Belgium | 2018 | 2012–2015 | P | RCT | 62 | 9 | Metachronous; OP, RTx. | ≤3 | Industrial |
| Steuber, prostate | Six European and one US center | 2019 | 1993–2014 | R | PSM | 659 | 9 | Metachronous; OP & adjuvant RTx (biochemical failure) | ≤5 | None |
| Parker, prostate | 117 centers in UK and Swiss | 2018 | 2013–2016 | P | RCT | 819 | 9 | Synchronous | ≤3 (low burden subgroup) | Industrial and government |
| Tsumura, prostate | Kitasato University, Japan. | 2019 | 2003–2013 | R | N/A | 40 | 7 | Synchronous | ≤5 | None |
| Giessen, colorectal | 48 German centers | 2013 | 2000–2004 | P | more N-, better PS | 253 | 7 | Synchronous and metachronous; OP (95%) | 1 (~95% of patients) | Industrial |
| Ruers, colorectal | 22 European centers | 2017 | 2002–2007 | P | RCT | 119 | 9 | Synchronous and metachronous | ≤9, all resectable or ablatable | None |
| Ruo, colorectal | Memorial Sloan Kettering Cancer Center, US | 2003 | 1996–1999 | R | more comorbidity (p=0.04), more liver only and single mets. (p=0.02) | 230 | 7 | Synchronous | ≤3 | None |
| Palma, multiple | 10 institutions in Canada, Netherlands, Scotland, and Australia | 2019 | 2012–2016 | P | RCT | 99 | 9 | Metachronous; no progression after definitive Tx. | ≤5 | Industrial |
| Chen Y, esophagus | Wuhan, Zengzhou Univ, China | 2019 | 2012–2015 | R | no significant difference | 461 | 8 | Synchronous | ≤3 | None |
| Depypere, esophagus | University Hospitals Leuven, Belgium | 2018 | 2002–2015 | R | N/A | 20 | 7 | Synchronous or metachronous; NAC(R)T | 3–5 mets in single organ | None |
| Chen J, HCC | Sun Yat-sen University Cancer Center, China. | 2018 | 2013–2016 | R | PSM | 68 | 9 | Synchronous | ≤5 in lung | None |
| Pan, HCC | Sun Yat-sen University Cancer Center, China. | 2017 | 2004–2013 | R | PSM | 92 | 9 | Synchronous | N/A | None |
| Morino, bile duct | Kyoto University, Japan. | 2020 | 1996–2015 | R | PSM, more ICC (p<0.001), more local mets. location (p=0.005) | 67 | 8 | Metachronous; R0 or R1 resection | ≤3 | None |
| Schulz, head and neck | Klinikum rechts der Isar, Germany. | 2018 | 2001–2016 | R | intentioned match | 47 | 7 | Synchronous and metachronous; OP, CTx., RT | 1 (77%), but ranged up to 10 | None |
| Falk, sarcoma | 15 centers, France | 2015 | 2000–2012 | R | smaller primary tumor (p=0.04), more controlled primary (p=0.0003), less lung mets (p=0.006) | 281 | 7 | Synchronous and metachronous; OP 93%, R0 62% R1 23% | ≤5 | Industrial |

Abbreviations: NOS, Newcastle-Ottawa scale; NSCLC, non-small cell lung cancer; SCLC, small cell lung cancer; HCC, hepatocellular carcinoma; R, retrospective; N/A, not assessable; OP, operation; P, prospective; RCT, randomized controlled trial; PR, partial remission; SD, stable disease; CTx., chemotherapy; PSM, propensity score matching; TKI, tyrosine kinase inhibitor; PSA, prostate-specific antigen; RTx, radiotherapy; PS, performance status

**Figure 1.**
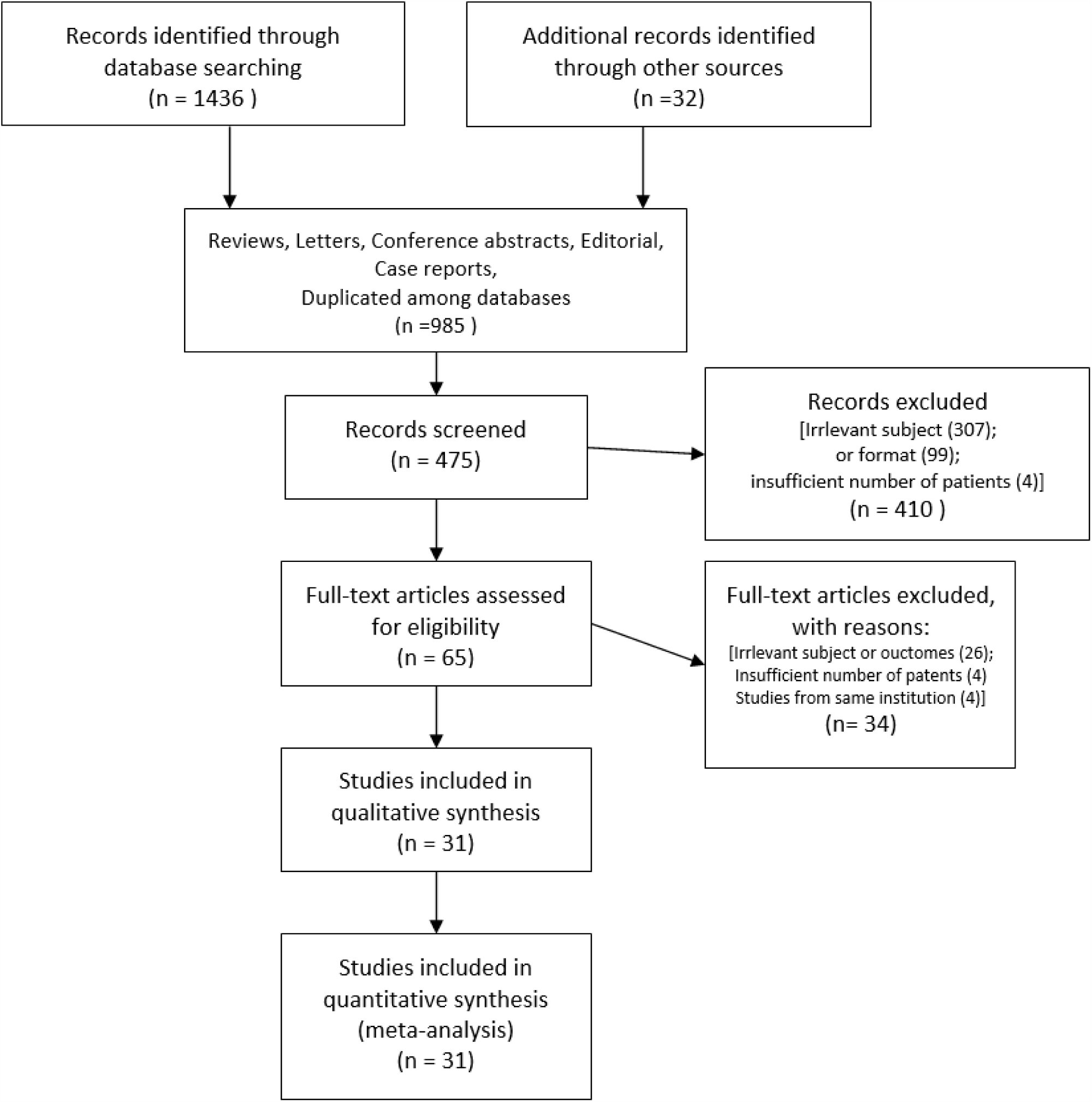
Study selection process.

LCT was performed principally to treat metastatic disease in 24 studies (77%) and primary malignant disease in nine. Radiotherapy was the LCT modality of choice in 22 studies (71%), while surgical resection was performed in 19 (61%). Radiofrequency or microwave ablation was used in few studies involving patients with liver neoplasms or metastases. Although only three studies reported statistically significant differences in the number of metastases between the study arms, 12 of the 22 studies (55%) reported a higher frequency of single or low-number metastases, without statistical significance, in the LCT arms. Clinical data from the studies are shown in Table 2.

**Table 2.** Clinical information of included studies

| Author, target disease | n | No. of oligomet s. | Site | Target of LCT | Modality of LCT | n | No. of oligomet s. | Site | Control | Median FU | OS (LCT arm vs. control arm) |  |  | PFS (LCT arm vs. control arm) |  |  |
| --- | --- | --- | --- | --- | --- | --- | --- | --- | --- | --- | --- | --- | --- | --- | --- | --- |
|  |  |  |  |  |  |  |  |  |  |  | Median (months) | 1/2 year rate | p | Median (months) | 1/2 year rate | p |
|  |  |  | LCT arm |  |  |  |  | Control arm |  |  |  |  |  |  |  |  |
| He, NSCLC | 11 | 1 (60%);<br>2 (40%) | Lung 100% | M | resection of pulmonary mets and/or adj. CTx. | 10 | N/A | Lung 100% | CTx. | 37.5 | 37 vs. 11.6 | 100/70% vs. 80/40% | 0.026 |  |  |  |
| Iyengar, NSCLC | 14 | 2 (50%);<br>3-4 (28.6%) | Lung or mediastinum >70% | M | SBRT & CTx. | 15 | 2 (40%);<br>3-4(33%) | Lung or mediastinum >70% | CTx. | 9.6 | not reached |  |  | 9.7 vs. 3.5 | 1yr: 35.7% vs 13.3% | 0.01 |
| Sheu, NSCLC | 60 | mean 1.28 | Brain (~50%) | M and P | conventional RTx. (76%) | 14 | mean 1.23 | Brain (~50%) | CTx. |  |  | 83.3/58.3 vs. 35.7/0% | <0.01 |  | 1yr: 46.7% vs. 18.2% | <0.01 |
| Yano, NSCLC | 44 |  |  | M (recurrence) | surgery or RTx. And/or CTx. | 49 |  |  | CTx. or SOC | ~4 year | 74 vs 10.9 | 77.3/61.4% vs. 46.9/24.5% | <0.05 |  |  |  |
| Frost, NSCLC | 90 | 1 (85%);<br>2 (8%) | Brain 57%;<br>bone 10%;<br>lung 9% | M and/or P | Lobectomy, CCRT, SBRT; 79% received CTx. | 90 | 1 (76%);<br>2 (14%) | Brain 32%;<br>bone 22%;<br>lung 21% | CTx. (96%) | 32 vs. 19 | 60.4 vs 22.5 | 92.2/76 vs. 81.9/45.9% | <0.001 | 25.1 vs. 8.2 | 67.8/52.2 % vs. 31/8.9% | <0.001 |
| Gomez, NSCLC | 25 | 0-1 (68%);<br>2-3 (32%) | Brain 28%;<br>other 72% | M and/or P | RTx. or surgery & standard maintenance | 24 | 0-1 (62%);<br>2-3 (38%) | Brain 25%;<br>other 75% | Standard maintenance | 38.8 | 41.2 vs. 17 | 84/68% vs. 62.5/45.8% | 0.017 | 14.2 vs. 4.4 | 52/28% vs. 20.8/12.5 % | 0.022 |
| Gray, NSCLC | 38 | 1 (50%);<br>2-4 (50%) | Brain 100% | P | ATT (surgery or >45Gy RTx. to thorax); brain RTx. & CTx | 28 | 1 (50%);<br>2-4 (50%) | Brain 100% | CTx and/or Brain RTx. |  | 26.4 vs. 10.5 | 71/54% vs. 46/26% | <0.001 |  |  |  |
| Hu, NSCLC | 143 | 1-3 (81%);<br>4-5 (19%) | Brain 44%;<br>Bone 35% | M | surgery and/or radiotherapy and TKI | 88 | 1-3 (83%);<br>4-5 (17%) | Bone 42%;<br>lung 33% | CTx. (TKI) | 24 | 34 vs. 21 | 95.3/72.1% vs. 84.1/40.9% | 0.001 | 15 vs 10 | 60.7/18.6 % vs 33.3/10.8 % | <0.001 |
| Song, NSCLC | 35 | 1 (46%);<br>2 (29%);<br>3-5 (26%) | Lung 57%;<br>bone 40%;<br>liver 30% | M and/or P | surgery or RTx. and CTx. | 35 | 1 (23%);<br>2 (40%);<br>3-5 (37%) | Lung 60%;<br>bone 54% | CTx. |  |  | 51.4/28.6% vs. 31.4/5.7% | 0.002 |  |  |  |
| Xu Q, NSCLC | 51 | 1 (49%);<br>2-3 (51%) |  | P and/or M | surgery or RTx. After TKI CTx. | 39 | 1 (41%);<br>2-3 (51.3%) |  | CTx. (TKI) | 38 | 40.9 vs. 30.8 | 96.1/86.3% vs. 94.9/71.8% | <0.001 | 20.6 vs. 13.9 | 86.3/25.6 % vs. 70.5/0% | <0.001 |
| Ni, NSCLC | 34 | 1-3 (85%); 4-5(15%) | Lung (40%); liver (23%); adrenal gland (16%) | M and/or P | TKI & MWA | 52 | 1-3 (89%); 4-5 (11%) | Lung (32%); bone (23%); liver (20%) | CTx. (TKI) | 36 | 34.8 vs. 22.7 | 94.1/67.6% vs. 90.3/46.2% | 0.04 | 16.7 vs. 12.9 | 88.2/23.5 % vs. 61.5/0% | 0.02 |
| Shang, NSCLC (postop) | 105 | 1 (73%); 2-5 (27%) | LN (46%); brain (24%); lung (19%) | M and/or P | RTx. or RFA and/or CTx. | 47 | 1 (72%); 2-5 (28%) | LN (72%) lung (32%) | CTx. or BSC | 19 | 19 vs. 20 | 1yr: 72.4 vs 72.3% | 0.519 | 10 vs. 7 | 1yr: 40.9 vs. 29.8% | 0.006 |
| Gore, SCLC (extended) | 44 | 1 (32%); 2-4 (68%) | Adrenal 25%; distant LN 23%; liver 23% | P | PCI and cRTx. (45Gy/15F) | 42 | 1 (41%); 2-4 (60%) | Distant LN 31%; Bone 26%; Liver 24% | PCI | 9 | 13.8 vs. 15.8 | 1yr: 50.8 vs. 60.1% | 0.21 | 4.9 vs. 2.9 | 1yr: 23.9 vs. 20.5% | 0.01 |
| Xu SCLC (extended) | 22 |  |  | M and/or P | CTx and RTx | 22 |  |  | CTx. | 36.4 |  | 72.7/25.2 vs. 18.2/12.7% | 0.002 |  | 40.9/19.3 vs. 9.1/4.8% | 0.006 |
| Bouman-Wammes, prostate | 43 | 1 (81%); 2 (14%) | LN 77%; bone 21% | M | SBRT (mostly 30Gy/3F or 35Gy/7F) | 20 | 1 (45%); 2 (40%) | LN 65%; Bone 35% | Active surveillance |  |  |  |  | 17.3 vs 4.2 | 72.1/35.8 % vs. 22.6/0% | <0.001 |
| Lan, prostate | 35 | 1 (26%); 2 (37%); 3 (20%) | Bone 100% | P | Prostatectomy & ADT | 76 | 1(8%); 2(32%); 3(30%) | Bone 100% | ADT | 35 |  | CSS 3/5yr: 90.8/63.6% vs. 87.9/74.9% | 0.773 | (PSA-RFS) 32 vs. 17 | 82.8/62.8 % vs. 65.8/38.2 % | 0.184 |
| Ost, prostate | 31 | 1 (58%); 2(19%); 3(22%) | LN 55%; non-nodal 45% | M | SBRT(81%) or resection | 31 | 1 (29%); 2 (32%); 3(39%) | LN 55%; non-nodal 45% | Active surveillance | 3 year |  |  |  | (ADT-free survival) 21 vs. 13 | 70.9/45.2% vs. 64.5/32.3% | 0.11 |
| Steuber, prostate | 165 |  | Pelvic LN ~90% | M | PLND or SBRT (≥30Gy/6F) and ADT | 494 |  | Pelvic LN ~90% | ADT |  |  | CSS 5/10 yr: 98.6/95.6 vs. 95.7/84.8% OS 3/5 yr: 99.2/98.7 vs. 98.2/95.4% | 0.03; 0.23 |  |  |  |
| Parker, prostate | 410 |  | Bone 76%; distant LN 36% | P | RT and ADT | 409 |  | Bone 76%; distant LN 34% | ADT | 37 |  | 98.8/92.5/82.6 % vs. 96.7/87.7/74.8 % | 0.007 |  | 89.6/72.8 % vs. 86.3/69.3 % | 0.033 |
| Tsumura, prostate | 22 |  | Bone or pelvic LN | M | metastatic RTx., prostate brachy & HTx. | 18 |  | Bone or pelvic LN | prostate brachy& HTx. | 62.5 |  |  |  |  | 94.4/88.9% vs. 95.5/73.3% | 0.027 |
| Giessen, colorectal | 38 | 1 (95%) | Liver 100% | M | Hepatic resection and CTx. | 215 | 1 (100%) | Liver 100% | CTx. |  | 48.0 (95% CI: 42-54) vs. 17.0 (95% CI: 13.9-20.1) | 97.4/89.5% vs. 68/37.6% | <0.001 | 16.6 vs. 6.5 | 63.2/36.8 % vs. 21.2/5.2% | <0.001 |
| Ruer, colorectal | 60 | 1-3 (48%); 4-6 (30%); 7-9 (22%) | Liver 100% | M | RFA, surgery and/or CTx. | 59 | 1-3 (31%); 4-6 (46%); 7-9 (24%) | Liver 100% | CTx. | 9.7 years | 45.6 vs 40.5 | 91.7/75% vs. 89.8/74.5% | 0.01 | 16.8 vs. 9.9 | 58.3/35% vs. 40.7/20.3% | 0.005 |
| Ruo, colorectal | 127 | 1 (68%); 2(26%); 3(6%) | Liver 56% | P | bowel surgery and CTx. | 103 | 1 (53%); 2(30%); 3(17%) | Liver 41% | CTx. (83.5%) |  | 16 vs. 9 | 63.8/25% vs. 35.9/6% | <0.001 |  |  |  |
| Palma, multiple | 66 | 1(46%); 2(29%); 3(18%) | lung 43%; bone 35% | M | SBRT and/or standard CTx. | 33 | 1 (36%); 2(40%); 3(18%) | Lung 53%; bone 31% | CTx. | 26 vs. 25 | 41 vs. 28 | 84.3/69.7% vs. 87.4/60.6% | 0.09 | 12 vs. 6 | 54.5/36.4% vs. 22.7/15.2% | 0.001 |
| Chen Y, esophagus | 196 |  |  | M and P | CCRT (IMRT, 50Gy/25F to primary; 45Gy/15F to metastases; cisplatin/paclitaxel) | 265 |  |  | CTx | 11.5 | 16.8 (95% CI: 15.5-18.1) vs 14.8 (95% CI: 13.2-16/4) | 72.8/27.2% vs. 63.5/17.5% | 0.056 | 8.7 vs. 7.3 | 27.6/4.7% vs. 21.9/0.9% | 0.002 |
| Depypere, esophagus | 10 |  | Lung 50%; adrenal 20% | P | esophagectomy +/- lung metastatectomy | 10 |  | Liver 50%; brain 30% | CTx. |  | 21.4 vs 12.1 | 80/40% vs. 50/10% | 0.042 |  |  |  |
| Chen J, HCC | 34 |  | Lung 100% | M and/or P | TACE, RFA, resection & sorafenib | 34 |  | Lung 100% | Sorafenib | 8.4 | 18.4 vs. 7.4 | 67.6/47% vs. 35.3/23.5% | 0.015 | TTP: 3.1 vs. 2.3 | (TTP) 11.8/0% vs. 0/0% | 0.009 |
| Pan, HCC | 46 | Mean 2.22 +/- 1.35 | LN 100% | M (lymph node) | RFA; and BSC or sorafenib | 46 | Mean 2.74 +/- 1.37 | LN 100% | BSC or sorafenib | 14 vs 13.8 | 13 vs. 7.8 | 58.3%/11.7% vs. 17.9/0% | 0.001 |  |  |  |
| Morino, bile duct | 33 | Median 1 (1-3) | Liver 39%; LN 27%; lung 12% | M (recurrence) | Surgery, RT, RFA, TACE and/or CTx. | 34 | Median 1 (1-3) | Local 35.3%; liver 29%; LN 20.5% | CTx. or BSC | 12.6 | 48.6 vs. 14.2 | 97/84.8% vs. 64.7/20.5% | <0.001 |  |  |  |
| Schulz, head and neck | 37 | 1 (70%); 2-3 (16%) | Lung 59%; bone 22% | M | RTx. or resection and/or CTx. | 10 | 1 (100%) | Lung 90% | CTx. or BSC |  | 24 vs. 7 | 67.6%/51.3% vs. 20%/10% | NA |  |  |  |
| Falk, sarcoma | 164 |  | Lung 51%; liver 7% | M | RTx. (>50Gy), RFA, OP removing all mets +/- CTx. | 117 |  | Lung 69%; liver 7% | CTx. in majority | 25.7 |  | 79.6/63.6% vs. 52.3/36.3% | <0.0001 |  |  |  |

Abbreviations: LCT, local consolidation therapy; OS, overall survival; PFS, progression free survival; CTx., chemotherapy; M, metastases; P, primary disease; NSCLC, non-small cell lung cancer; RTx., radiotherapy; CCRT, concurrent chemoradiotherapy; SBRT, stereotactic body radiotherapy; ATT, aggressive thoracic therapy; TKI, tyrosine kinase inhibitor; MWA, microwave ablation; SCLC, small cell lung cancer; RFA, radiofrequency ablation; LN, lymph node; BSC, best supportive care; PCI, prophylactic cranial irradiation; ADT, androgen deprivation therapy; PLND, pelvic lymph node dissection; IMRT, intensity modulated radiotherapy; TACE, transarterial chemoradiotherapy; TTP, time to progression; OP, operation

In pooled analysis of OS, the odds ratios (ORs) were 3.04 (95% confidence interval [CI]: 2.28–4.06, p<0.001), 2.56 (95% CI: 1.79–3.66, p<0.001), and 1.41 (95% CI: 1.02–1.95, p=0.041) for all studies, balanced studies, and RCTs, respectively. On analyses of PFS, the pooled ORs were 2.82 (95% CI: 1.96–4.06, p<0.001), 2.32 (95% CI: 1.60–3.38, p<0.001), and 1.39 (95% CI: 1.09–1.80, p=0.009) among all studies, balanced studies, and RCTs, respectively. The pooled ORs for OS among studies principally investigating metastases and primary diseases were 3.34 (95% CI: 2.40–4.66, p<0.001) and 2.22 (95% CI: 1.21–4.08, p=0.010), respectively, with no significant difference in subgroup comparisons (p=0.248); the corresponding ORs for PFS were 3.34 (95% CI: 2.18-5.13) and 1.60 (95% CI: 0.99-2.59), respectively, with a significant difference between subgroups (p=0.025). Heterogeneity was significant in most pooled analyses, but was low and insignificant in pooled analyses of RCTs alone. Possible publication biases were noted in the pooled OS analyses of all studies and those only investigating metastases, as well as in pooled PFS analyses of all studies, balanced studies, and studies investigating metastases. The main results are depicted as forest plots in Figure 2, and detailed pooled analysis results are shown in Table 3.

**Table 3.**
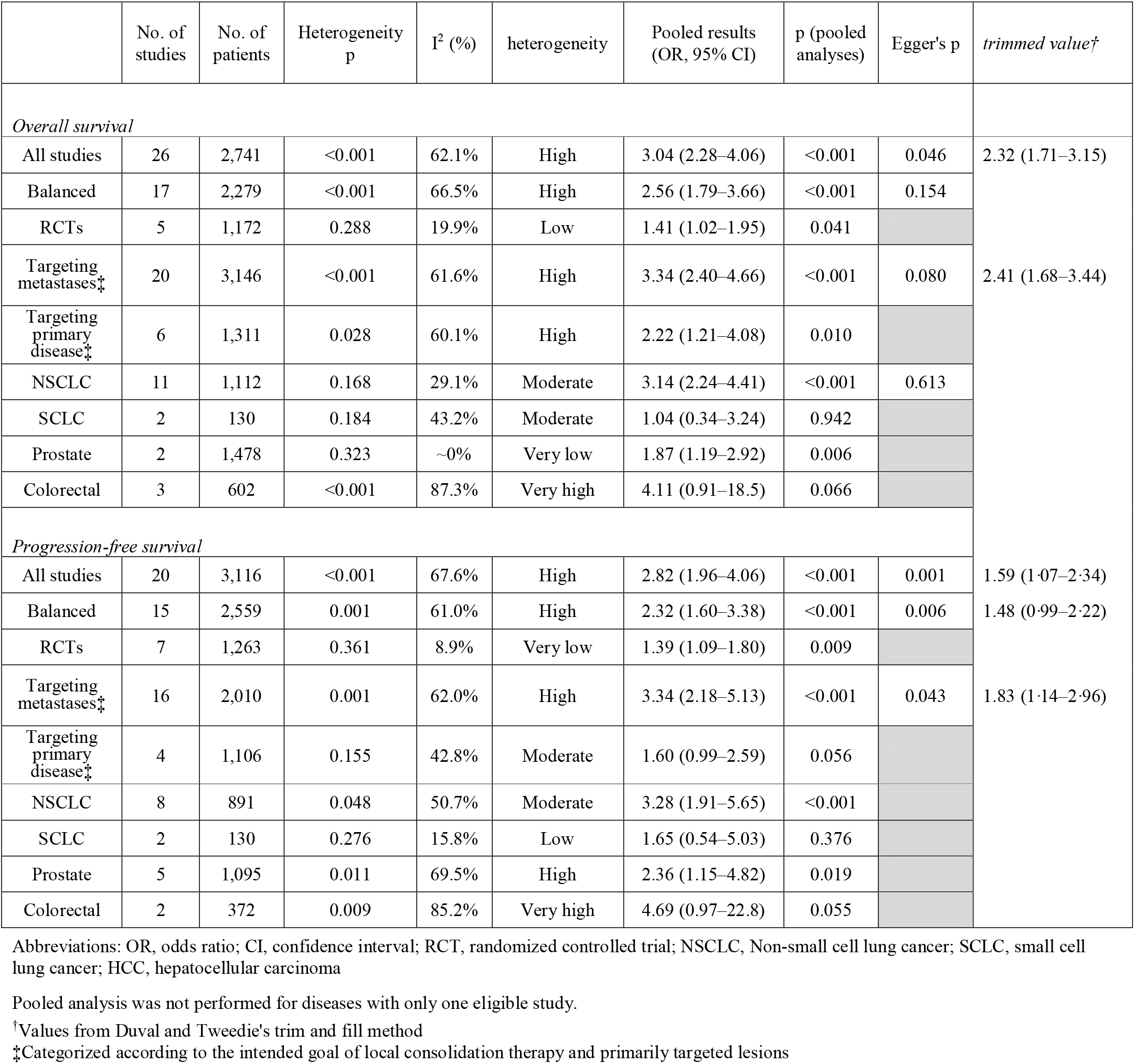
Pooled results of endpoints

**Figure 2.**
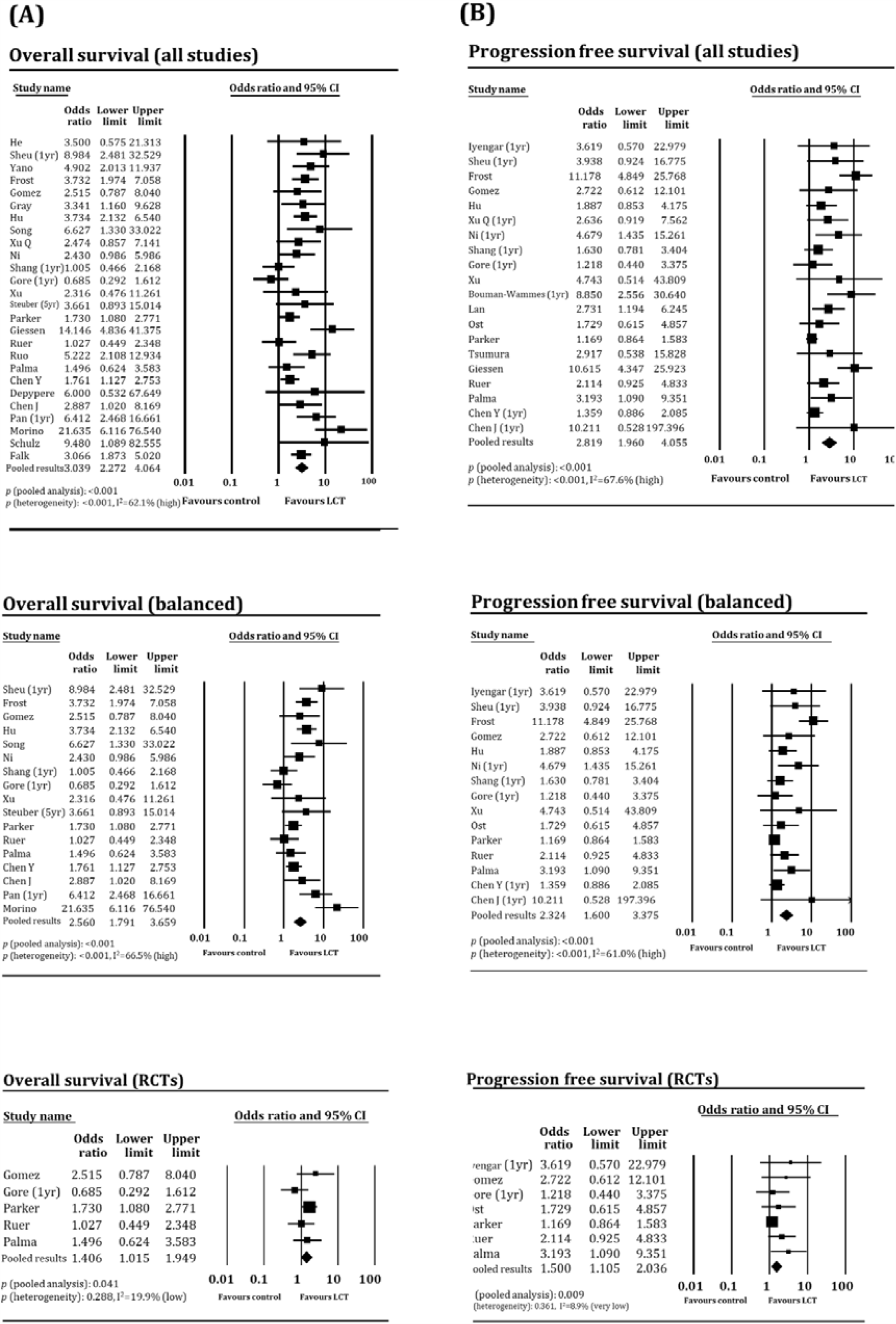
Forest plots of pooled analyses of (A) overall survival per all, balanced, and randomized controlled trials and (B) progression-free survival per all, balanced, and randomized controlled trials.

In pooled analyses of OS according to cancer types, the benefit of LCT was higher in patients with NSCLC (OR: 3.14, p<0.001; pooled two-year OS: 65.2% vs. 37.0%) and colorectal cancer (OR: 4.11, p=0.066; two-year OS: 66.2% vs. 33.2%) than in those with prostate cancer (OR: 1.87, p=0.006; three-year OS: 95.6% vs. 92.6%) and SCLC (OR: 1.04, p=0.942; 60.7% vs. 42.8%). Heterogeneity was not significant in the pooled OS analyses of patients with NSCLC, SCLC, and prostate cancer, but was significant for those with colorectal cancer. The results were similar in pooled analyses of PFS; the benefit of LCT was higher for patients with NSCLC (OR: 3.28, p<0.001; pooled two-year PFS: 28.9% vs. 8.6%) and colorectal cancer (OR: 4.69, p=0.055; two-year PFS: 35.7% vs. 10.5%) and was lower for those with prostate cancer (OR: 2.36, p=0.019, two-year PFS: 82.7% vs. 61.7%), and SCLC (OR: 1.65, p=0.376; one-year PFS: 30.9% vs. 16.6%). Heterogeneity was not significant in pooled PFS analyses of patients with NSCLC and SCLC but was significant for those with prostate and colorectal cancers. Detailed results according to disease type are shown in Tables 3 and 4.

**Table 4.**
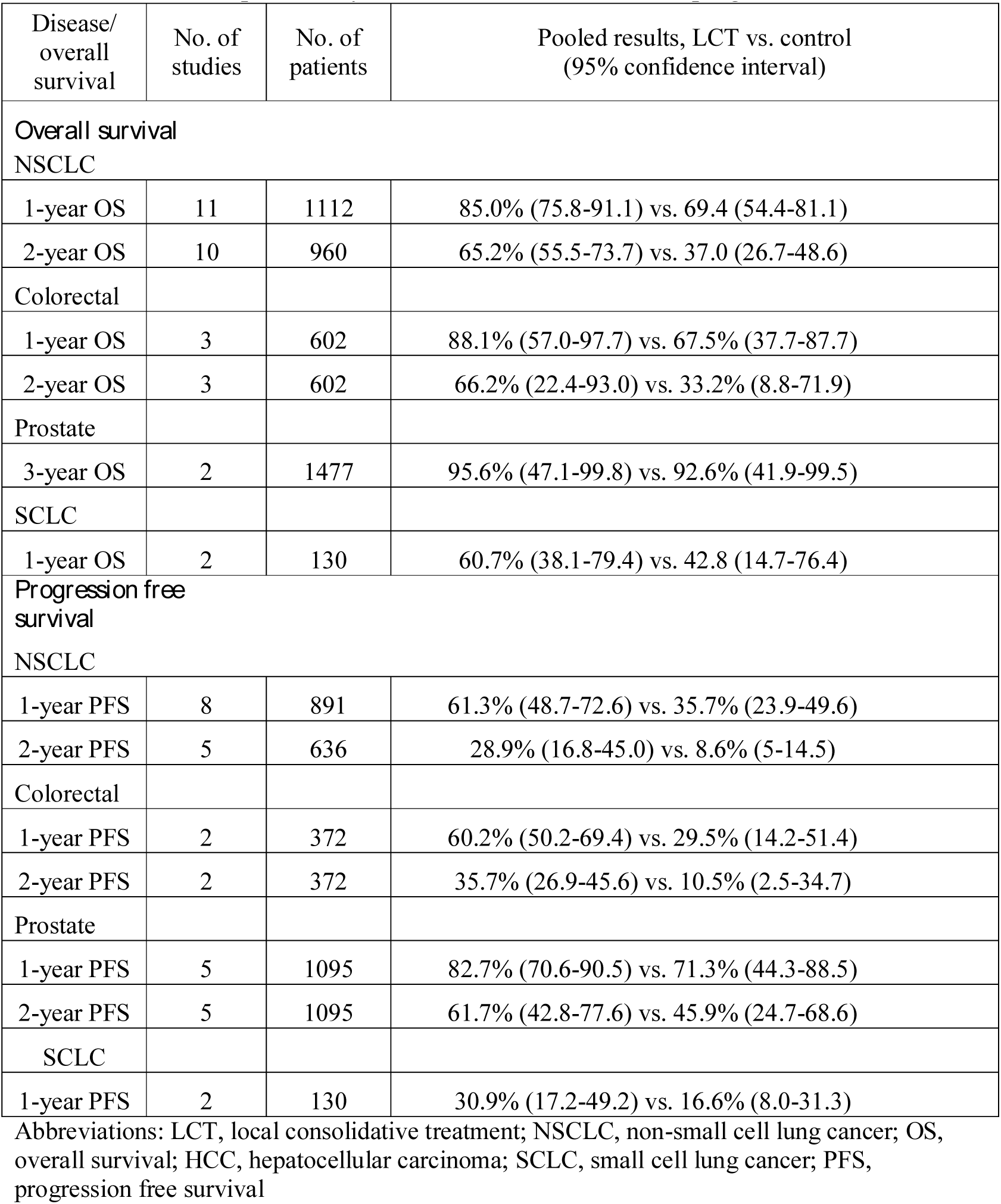
Pooled temporal analyses of numerical overall- and progression free survival

| Disease/<br>overall<br>survival | No. of<br>studies | No. of<br>patients | Pooled results, LCT vs. control<br>(95% confidence interval) |
| --- | --- | --- | --- |
| *Overall survival* |  |  |  |
| NSCLC |  |  |  |
| 1-year OS | 11 | 1112 | 85.0% (75.8-91.1) vs. 69.4 (54.4-81.1) |
| 2-year OS | 10 | 960 | 65.2% (55.5-73.7) vs. 37.0 (26.7-48.6) |
| Colorectal |  |  |  |
| 1-year OS | 3 | 602 | 88.1% (57.0-97.7) vs. 67.5% (37.7-87.7) |
| 2-year OS | 3 | 602 | 66.2% (22.4-93.0) vs. 33.2% (8.8-71.9) |
| Prostate |  |  |  |
| 3-year OS | 2 | 1477 | 95.6% (47.1-99.8) vs. 92.6% (41.9-99.5) |
| SCLC |  |  |  |
| 1-year OS | 2 | 130 | 60.7% (38.1-79.4) vs. 42.8 (14.7-76.4) |
| *Progression free survival* |  |  |  |
| NSCLC |  |  |  |
| 1-year PFS | 8 | 891 | 61.3% (48.7-72.6) vs. 35.7% (23.9-49.6) |
| 2-year PFS | 5 | 636 | 28.9% (16.8-45.0) vs. 8.6% (5-14.5) |
| Colorectal |  |  |  |
| 1-year PFS | 2 | 372 | 60.2% (50.2-69.4) vs. 29.5% (14.2-51.4) |
| 2-year PFS | 2 | 372 | 35.7% (26.9-45.6) vs. 10.5% (2.5-34.7) |
| Prostate |  |  |  |
| 1-year PFS | 5 | 1095 | 82.7% (70.6-90.5) vs. 71.3% (44.3-88.5) |
| 2-year PFS | 5 | 1095 | 61.7% (42.8-77.6) vs. 45.9% (24.7-68.6) |
| SCLC |  |  |  |
| 1-year PFS | 2 | 130 | 30.9% (17.2-49.2) vs. 16.6% (8.0-31.3) |

Abbreviations: LCT, local consolidative treatment; NSCLC, non-small cell lung cancer; OS, overall survival; HCC, hepatocellular carcinoma; SCLC, small cell lung cancer; PFS, progression free survival

Twelve of 31 studies (38.7%) involving 2,176 patients included data of complications related to treatment modalities. Palma et al. (21) reported three grade 5 cases (4.5%) possibly related to SBRT, whereas Gore et al. (27) reported a significantly higher rate of grade 3 toxicity (24.8% vs. 9.5%) in the LCT arm (with one patient developing grade 5 toxicity). Ruo et al. (36) reported a postoperative serious morbidity rate of 20.5%, with two patients developing grade 5 complications within 30 days of elective colorectal surgery. Ni et al. (9) reported that 9.3% of patients needed chest tube insertion, while no serious toxicities were reported in the control arm. Otherwise, no significant additional toxicity due to LCTs were reported in eight studies in which LCT consisted mainly of radiotherapy (Table 5).

**Table 5.** Assessment of complications

| Author, target disease | Modality of LCT | n | Control | n | Grade $\geq 3$ toxicity |
| --- | --- | --- | --- | --- | --- |
| Iyengar, NSCLC | SBRT & CTx. | 14 | CTx. | 15 | A total of 7 (50%) and 9 (60%) cases for LCT and control, respectively; no G5 toxicity |
| Gomez, NSCLC | RT or surgery & standard maintenance | 25 | Standard maintenance | 24 | 2 cases with G3 esophagitis in LCT; 1 G3 fatigue and 1 G3 anemia in control |
| Ni, NSCLC | TKI & MWA | 34 | TKI | 52 | 4 (9.3%) of MWA group needed chest tube drainage; no G$\geq 3$ toxicity related to TKI |
| Shang, NSCLC (postop) | RT or RFA and/or CTx. | 105 | CTx. or BSC | 47 | Overall: 24.8% vs. 21.2% (m/c Cx.: myelosuppression)<br>1 case (0.9%) of grade 5 (infection) in LCT arm |
| Gore, SCLC | PCI and cRT (45 Gy/15 F) | 44 | PCI | 42 | Overall: 25% vs. 9.5%; 1 case of G5 pneumonitis in LCT arm |
| Bouman-Wammes, prostate | SBRT (mostly 30Gy/3F or 35Gy/7F) | 43 | Active surveillance | 20 | No SBRT-related toxicity |
| Ost, prostate | SBRT (81%) or resection | 31 | Active surveillance | 31 | No grade 2 or higher toxicity in LCT arm |
| Parker, prostate | RT and ADT | 410 | ADT | 409 | No data in low metastatic burden subgroup; 4% vs 1% for whole population |
| Tsumura, prostate | RT to metastases, prostate brachytherapy & HTx. | 22 | prostate brachytherapy & HTx. | 18 | No difference in grade $\geq 2$ toxicity |
| Ruo, colorectal | Bowel surgery and CTx. | 127 | CTx. (83.5%) | 103 | Grade 5: 2 cases (1.6%); postop OP morbidity (20.5%) |
| Palma, multiple | SBRT and/or standard CTx. | 66 | CTx. | 33 | Higher rate in LCT (10.6% vs. 3%); 3 grade 5 cases due to SBRT |
| Chen Y, esophagus | CCRT (IMRT, 50 Gy/25 F to primary; 45 Gy/15 F to metastases; cisplatin/paclitaxel) | 196 | CTx | 265 | No significant difference between arms |

Abbreviations: LCT, local consolidation therapy; NSCLC, non-small cell lung cancer; SBRT, stereotactic body radiotherapy; CTx., chemotherapy; RT, radiotherapy; TKI, tyrosine kinase inhibitor; MWA, microwave ablation; BSC, best supportive care; PCI, prophylactic cranial irradiation; SCLC, small cell lung cancer; cRT, chest radiotherapy; ADT, androgen deprivation therapy; HTx, hormone therapy; OP, operation; CCRT, concurrent chemoradiation; IMRT, intensity-modulated radiotherapy.

## Discussion

The concept of *oligometastases* has attracted significant interest as a potential curative opportunity for patients whose diseases were deemed intractable. Nevertheless, the benefit of LCT and the existence of an “intermediate stage” remains controversial. Molecular studies to identify disease-specific biomarkers or gene profiles have shown promising results (42, 43); however, external or internal validation were lacking or unsuccessful.(10) Clinical data reported to date are extremely heterogeneous, making it difficult for physicians to decide whether to apply LCTs. Currently, practical decisions for the application of LCTs are mostly based on single-arm studies that demonstrated favorable survival outcomes in selected patients. However, complications from LCTs (possibility of missed occult metastases) as well as the distribution of economic and medical resources are all issues to consider.(6, 9)

The present meta-analysis successfully demonstrated the clinical significance of the “oligometastasis” status, as patients with this status can benefit from LCTs. Regarding OS, the pooled results from all studies (OR: 3.04, p<0.001) and balanced studies (e.g., those without significant differences in major clinical indicators; OR: 2.56, p<0.001) were significant, with a high degree of heterogeneity. Possible publication biases were noted, and the trimmed value after sensitivity analysis was lower than the original value (OR: 2.32). The OR was also significant on the pooled analysis of RCTs (OR: 1.41, p=0.041) with a low degree of heterogeneity, but was lower in magnitude than the ORs of the total and balanced studies. The pooled PFS results also showed trends similar to OS. The statistically significant results (with low heterogeneity) obtained from the pooled analyses of RCTs, regarding both OS and PFS, supporting the existence of the clinical concept of oligometastases that can be treated with LCTs. However, the extent of successful oligometastasis treatment might be smaller than described in literature reporting data from observational studies, which showed more favorable survival outcomes than expected.(44) The significant heterogeneity and possible publication biases additionally indicate that selection biases might be present in the literature despite statistical efforts to balance both arms. For example, patients in the LCT arms of 12 of 22 available studies (55%) tended to have fewer numbers of metastases, although the differences were not statistically significant.

The majority of the clinical literature around oligometastases is disease-specific, and few studies have compared outcomes between different cancer types. According to subgroup analyses based on cancer types, the benefits of LCT as well as survival outcomes vary widely among disease entities. The survival benefits from LCTs were most prominent for patients with NSCLC and colorectal cancers, which are the most vigorously researched diseases to date.(9) Although the oncologic benefit of LCT was also significant among patients with prostate cancer, the extent of this benefit was relatively smaller. Survival outcomes of patients with oligometastatic prostate cancer was favorable regardless of application of LCTs, suggesting that less aggressive tumor biology of prostate cancer than other cancer types (e.g. NSCLC or colorectal cancer). The pooled OR for patients with SCLC did not show significance for either OS or PFS (p=0.942 and 0.376, respectively), suggesting a lower influence of LCTs on oncologic outcomes. This was consistent with the conventional notion that SCLC behaves more like a systemic disease and metastasizes early.(45) Little is known about whether LCT that targets the primary disease is as beneficial as that which targets all the oligometastatic foci; other than for nephrectomy and metastatic renal cell carcinomas, data regarding its benefit are mostly preclinical or exploratory.(46) Although the OS benefit was not significantly different in subgroup comparisons, the PFS benefit differed between studies investigating primary diseases versus those examining metastases. Hence, our subgroup analyses suggest that applying LCTs to primary malignant diseases also produces oncologic benefits, although to lesser degrees. The meta-analysis methodology is limited in its ability to evaluate the causes of the abovementioned differences. However, our results will help with clinical decision-making in practice, and will provide hypotheses for future oligometastases research to identify differences between cancer types and define additional LCT targets.

Although the majority of studies revealed no excessive complications, the unconditional application of LCT might not be justified because several investigators reported additional toxicities including few grade 5 complications.(9, 21, 27, 36) That LCTs showed benefits of lesser magnitudes in pooled analyses of RCTs than in overall pooled analyses suggests that a conservative approach is necessary when applying LCTs for oligometastatic diseases. From a practical perspective, the efficacy and feasibility of LCTs (as well as systemic treatments) should be discussed in a multidisciplinary setting. Radiotherapy should be applied when metastatic foci can be encompassed in the target field, and surgery should be considered when the complete resection of all metastatic lesions is feasible. The clinical conditions of the patients, as well as the type of cancers that can successfully be treated with the application of LCTs, should also be considered.

We included studies with multiple cancer types, which is not an uncommon approach in studies investigating LCTs for oligometastases.(47) The inherent heterogeneities might be criticized using the famous metaphoric phrase “*combining apples and oranges*.” According to Borenstein et al.,(48) a good meta-analysis aims to *synthesize* rather than simply report firm pooled effect sizes, and strives to explain phenomena that aid in clinical decision-making in practice. In other words, rather than avoiding heterogeneity, it should to be statistically assessed and clinically interpreted. Although controversial, meta-analyses including non-RCTs with sound statistical methods can shed light on the knowledge gaps in clinical practice and literature that cannot be addressed with RCTs.(49, 50) Therefore, our study might be one of the best available tools to evaluate the literature pool and assist in clinical decision-making, thereby adopting Dr. Rosenthal’s response to the abovementioned metaphoric phrase: “*It makes sense if your goal is to produce a fruit salad*.” (48) Other limitations include that, the small number of studies and patients in trials other than those involving NSCLC, prostate cancer, and colorectal cancer, and methodological limitations of meta-analysis that can only interpret the outcomes and cannot determine the cause.

## Conclusion

Our study demonstrates the oncologic benefits of LCTs in oligometastatic setting, regarding both OS and PFS. Although the benefits were also observed when analyzing RCTs, their extent were smaller than expected from literature data that included observational studies. LCT benefits were more prominent against oligometastases from NSCLC and colorectal cancer. Complications due to modern LCTs were generally low, although efforts to minimize toxicities are still necessary. Therefore, the appropriate LCTs should be selected carefully considering their efficacy and feasibility while also noting systemic treatments, clinical conditions, and disease types.

## Data Availability

N/A

## Ethical consideration and consent for publication

Ethical approval was not required because this study retrieved and synthesized data that are already published.

## Authors’ contributions

CHR did the conceptualization, writing-original draft & editing, data curation; ISS did the statistical analysis as a biostatistician; SP did the data curation & recruiting; HYL did the supervision. All authors read and approved the final manuscript

## Acknowledgements

This study was supported by the National Research Fund of Korea (NRF-2019M2D2A1A01031560).

